# Impact of circulating SARS-CoV-2 variants on mRNA vaccine-induced immunity in uninfected and previously infected individuals

**DOI:** 10.1101/2021.07.14.21260307

**Authors:** Carolina Lucas, Chantal B. F. Vogels, Inci Yildirim, Jessica E. Rothman, Peiwen Lu, Valter Monteiro, Jeff R. Gelhausen, Melissa Campbell, Julio Silva, Alexandra Tabachikova, M. Catherine Muenker, Mallery I. Breban, Joseph R. Fauver, Subhasis Mohanty, Jiefang Huang, Yale SARS-CoV-2 Genomic Surveillance Initiative, Claire Pearson, Anthony Muyombwe, Randy Downing, Jafar Razeq, Mary Petrone, Isabel Ott, Anne Watkins, Chaney Kalinich, Tara Alpert, Anderson Brito, Rebecca Earnest, Steven Murphy, Caleb Neal, Eva Laszlo, Ahmad Altajar, Irina Tikhonova, Christopher Castaldi, Shrikant Mane, Kaya Bilguvar, Nicholas Kerantzas, David Ferguson, Wade Schulz, Marie Landry, David Peaper, Albert C. Shaw, Albert I. Ko, Saad B. Omer, Nathan D. Grubaugh, Akiko Iwasaki

## Abstract

The emergence of SARS-CoV-2 variants with mutations in major neutralizing antibody-binding sites can affect humoral immunity induced by infection or vaccination^1–6^. We analysed the development of anti-SARS-CoV-2 antibody and T cell responses in previously infected (recovered) or uninfected (naive) individuals that received mRNA vaccines to SARS-CoV-2. While previously infected individuals sustained higher antibody titers than uninfected individuals post-vaccination, the latter reached comparable levels of neutralization responses to the ancestral strain than previously infected individuals 7 days after the second vaccine dose. T cell activation markers measured upon spike or nucleocapsid peptide in vitro stimulation showed a progressive increase after vaccination in the time-points analysed. Comprehensive analysis of plasma neutralization using 16 authentic isolates of distinct locally circulating SARS-CoV-2 variants revealed a range of reduction in the neutralization capacity associated with specific mutations in the spike gene: lineages with E484K and N501Y/T (e.g., B.1.351 and P.1) had the greatest reduction, followed by lineages with L452R (e.g., B.1.617.2) or with E484K (without N501Y/T). While both groups retained neutralization capacity against all variants, plasma from previously infected vaccinated individuals displayed overall better neutralization capacity when compared to plasma from uninfected individuals that also received two vaccine doses, pointing to vaccine boosters as a relevant future strategy to alleviate the impact of emerging variants on antibody neutralizing activity.

## Main

The ongoing evolution and emergence of severe acute respiratory syndrome coronavirus 2 (SARS-CoV-2) variants raise concerns about the effectiveness of monoclonal antibodies (mAbs) therapies and vaccines. The mRNA-based vaccines Pfizer-BioNTech BNT162b2 and Moderna mRNA-1273 encode a stabilized full-length SARS-CoV-2 spike ectodomain derived from the Wuhan-Hu-1 genetic sequence and elicit potent neutralizing antibodies (NAbs)^7,8^. However, emerging SARS-CoV-2 variants with mutations in the spike (S) gene, especially in NAb binding sites, have been associated with increased transmissibility^9,10^ as well as neutralization resistance to mAbs, convalescent plasma, and sera from vaccinated individuals^1–6^.

To better understand how immune responses triggered by SARS-CoV-2 infection, and/or vaccination, fare against emerging virus variants, we assembled a cohort of mRNA-vaccinated individuals, previously infected or not, and characterized virus-specific immunologic profiles. We examined the impact of SARS-CoV-2 variants containing many different key S gene mutations in mRNA-vaccinated individuals using a comprehensive set of full-length authentic SARS-CoV-2 isolates. Our variant panel included representatives that are currently classified as ‘variants of concern’ (lineages B.1.1.7 [Alpha], B.1.351 [Beta], P.1 [Gamma], and B.1.617.2 [Delta]), and ‘variants of interest’ (B.1.427/B.1.429 [Epsilon], B.1.525 [Eta], B.1.526 [Iota], and B.1.617.1 [Kappa]).

First, to characterize SARS-CoV-2-specific adaptive immune responses post mRNA COVID vaccines (Moderna or Pfizer), forty healthcare workers (HCWs) from the Yale-New Haven Hospital (YNHH), were enrolled in this study between November 2020 and January 2021, with a total of 198 samples. We stratified the vaccinated participants based on prior exposure to SARS-CoV-2 into previously infected (recovered) or uninfected (naive) groups. Previous infection was confirmed by RT–qPCR and SARS-CoV-2 IgG ELISA. The HCWs received mRNA vaccines, either Pfizer or Moderna, and we followed them longitudinally pre- and post-vaccination (Figure 1a). Cohort basic demographics, vaccination information, and serostatus are summarized in <u>Extended Data Table 1</u>. We collected plasma and peripheral blood mononuclear cells (PBMCs) sequentially in 5 time points covering a period of 98 days after the first vaccination dose. Samples were collected at baseline (prior to vaccination), 7- and 28- post first vaccination dose, and 7-, 28- and 70-days post second vaccination dose (Figure 1a). We determined antibody profiles, using both ELISA and neutralizations assays; and assessed cellular immune response, profiled by flow cytometry using frozen PBMCs.

**Figure 1.**
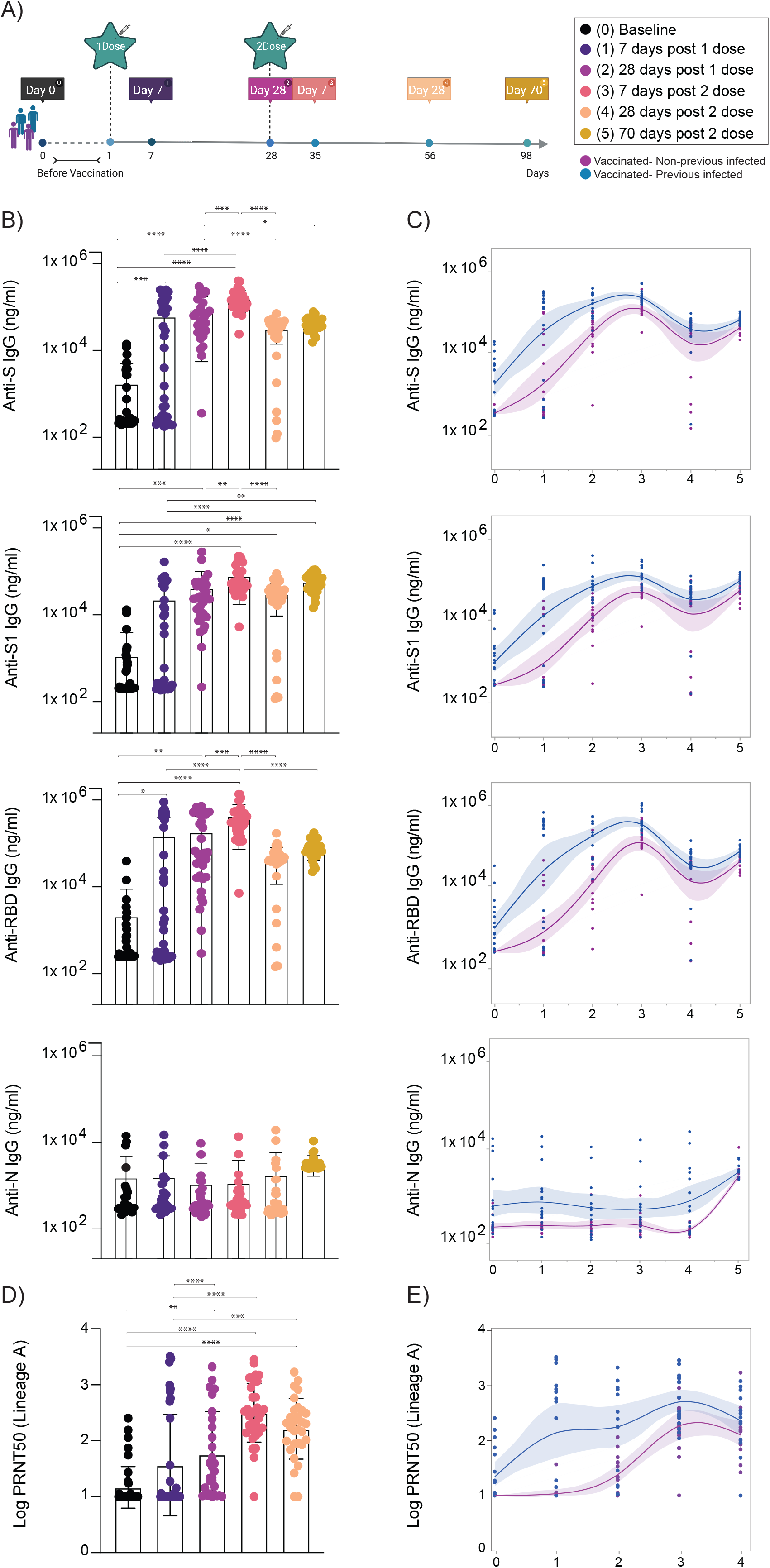
Temporal dynamics of anti-SARS-CoV-2 antibodies in vaccinated participants. **a**, Cohort timeline overview indicated by days post SARS-CoV-2 mRNA vaccination. HCW participants received 2 doses of the mRNA vaccine and plasma samples were collected as indicated. Baseline, prior to vaccination; Time point (TP) 1, 7 days post 1 dose; TP 2, 28 days post 1 dose; TP 3, 7 days post 2 dose; TP 4, 28 days post 2 dose; TP 5, 70 days post 2 dose. Participants were stratified based on previous exposure to SARS-CoV-2, indicated by purple (Vaccinated-uninfected) and blue (Vaccinated-Previously infected). **b, c**, Plasma reactivity to S protein, RBD and Nucleocapsid in vaccinated participants measured over time by ELISA. **a**, Anti-S, Anti-S1, Anti-RBD and Anti-N IgG levels. Significance was assessed by One-way ANOVA corrected for multiple comparisons using Tukey’s method. Boxes represent mean values ± standard deviations. **c**, Anti-S, Anti-S1, Anti-RBD and Anti-N IgG comparison in vaccinated participants previously infected or not to SARS-CoV-2. Longitudinal data plotted over time continuously. Regression lines are shown as blue (previously infected) and purple (uninfected). Lines indicate cross-sectional averages from each group with shading representing 95% CI and are coloured accordingly. TP, vaccination time point. Anti-S IgG (TP0, n = 37; TP1, n=35; TP2, n =30; TP3, n = 34; TP4, n=34; TP5, N=28). Anti-S1 IgG (TP0, n = 37; TP1, n=35; TP2, n =30; TP3, n = 34; TP4, n=34; TP5, N=27). Anti-RBD IgG (TP0, n = 37; TP1, n=35; TP2, n =30; TP3, n = 34; TP4, n=34; TP5, N=27). Anti-N IgG (TP0, n = 37; TP1, n=35; TP2, n =30; TP3, n = 34; TP4, n=34; TP5, N=27). S, spike. S1, spike subunit 1. RBD, receptor binding domain. N, nucleocapsid. Each dot represents a single individual. **d, e**, Longitudinal neutralization assay using wild-type SARS-CoV-2, ancestral strain (WA1, USA). **d**, Neutralization titer (PRNT50) over time. Significance was assessed by One-way ANOVA corrected for multiple comparisons using Tukey’s method. Boxes represent mean values ± standard deviations. **e**, Plasma neutralization capacity between vaccinated participants previously infected or not to SARS-CoV-2. Longitudinal data plotted over time continuously. Regression lines are shown as blue (previously infected) and purple (uninfected). Lines indicate cross-sectional averages from each group with shading representing 95% CI and are coloured accordingly. TP, vaccination time point (TP0, n = 38; TP1, n=35; TP2, n =30; TP3, n = 34; TP4, n=31). Each dot represents a single individual. ****p < .0001 ***p < .001 **p < .01*p < .05.

We found that mRNA vaccines induced high titers of virus-specific antibodies that declined over time, as previously reported^6,11^ (Figure 1b, c). After the first vaccine dose, over 97% vaccinated participants developed virus-specific IgG titers, which increased to 100% after the second dose. IgG titers against the S protein, spike subunit 1 (S1), and receptor binding domain (RBD) peaked 7 days post second vaccination dose (Figure 1 b, c). No differences were observed in antibody levels between vaccinated participants of different sexes and after stratification by age (<u>Extended Data Figure 1a</u>). Consistent with previous reports^7,12^, we found that virus-specific IgG levels were significantly higher in the previously infected vaccinated group than the uninfected vaccinated group (<u>Extended Data Figure 1b and Figure 1c</u>). As expected, given the absence of sequences encoding nucleocapsid (N) antigens in the mRNA vaccines, anti-N antibody titers remain stable over time for previously infected vaccinated individuals, and were not affected by vaccination in both the uninfected and previously infected groups (Figure 1 b, c). We next assessed plasma neutralization activity longitudinally against an authentic SARS-CoV-2 strain USA-WA1/2020 (lineage A), with a similar S gene amino acid sequence as Wuhan-Hu-1 used for the mRNA vaccine design, by a 50% plaque-reduction neutralization (PRNT50) assay. Neutralization activity directly correlates with anti-S and anti-RBD antibody titers, also peaking at 7 days post second dose (Figure 1 d, e). However, both groups displayed similar neutralization titers against the lineage A virus isolate at the peak of response (Figure 1 d, e). Our data indicate that despite faster and more exuberant antibody responses to viral proteins by previously infected vaccinated than uninfected vaccinated individuals, vaccination led to overall similar levels of neutralizing antibodies (NAb) after the second dose.

A robust T cell response has also been linked to efficient protective immunity against SARS-CoV-2^13–15^. Hence, we next longitudinally analyzed S- and N-reactive T cells responses in vaccinated individuals. To detect low-frequency peptide-specific T cell populations, we first expanded antigenic–specific T cells by stimulating PBMCs from vaccinated individuals with S and N peptide pools *ex vivo* for 6 days, followed by restimulation with the same peptide pools and analysis of activation markers after 12 hours. To cover the entire S protein, two peptide pools were used (S-I and S-II), while a single peptide pool was used for the nucleocapsid stimulation. S-reactive CD4^+^ and CD8^+^ T cells increased over time following vaccination (Figure 2 a, b), as evidenced by an increase in cells expressing the activation markers CD38 and HLA-DR; no differences were observed between the previously infected and uninfected vaccinated groups. Consistent with previous reports^16,17^, S-reactive CD4^+^ T cell responses were comparable among full-length lineage A and P.1 virus isolates. In contrast, S-reactive CD8^+^ T cell responses were only observed to lineage A, and not P.1, 28 days post second vaccination dose, suggesting that S-specific CD8 T cell responses can be affected by the mutations within the S gene of SARS-CoV-2 variants (Figure 2b). As expected, N-reactive T cells induced after stimulation with a N-peptide pool derived from the lineage A virus isolate were primarily observed in the previously infected vaccinated individuals (Figure 2b). Unexpectedly, we also observed elevated N-reactive CD4 T cells in previously infected individuals at 28 days post second dose, paralleling general activation of CD4 T cells (Figure 2b and <u>Extended Data Figure 2a</u>). Moreover, we observed increased levels of activated CD4^+^ T, Tfh, and antibody-secreting cells 28 days post vaccination booster <u>(Extended Data Figure 2</u>). Thus, T cell responses in vaccinated individuals display similar dynamics as antibody responses.

**Figure 2.**
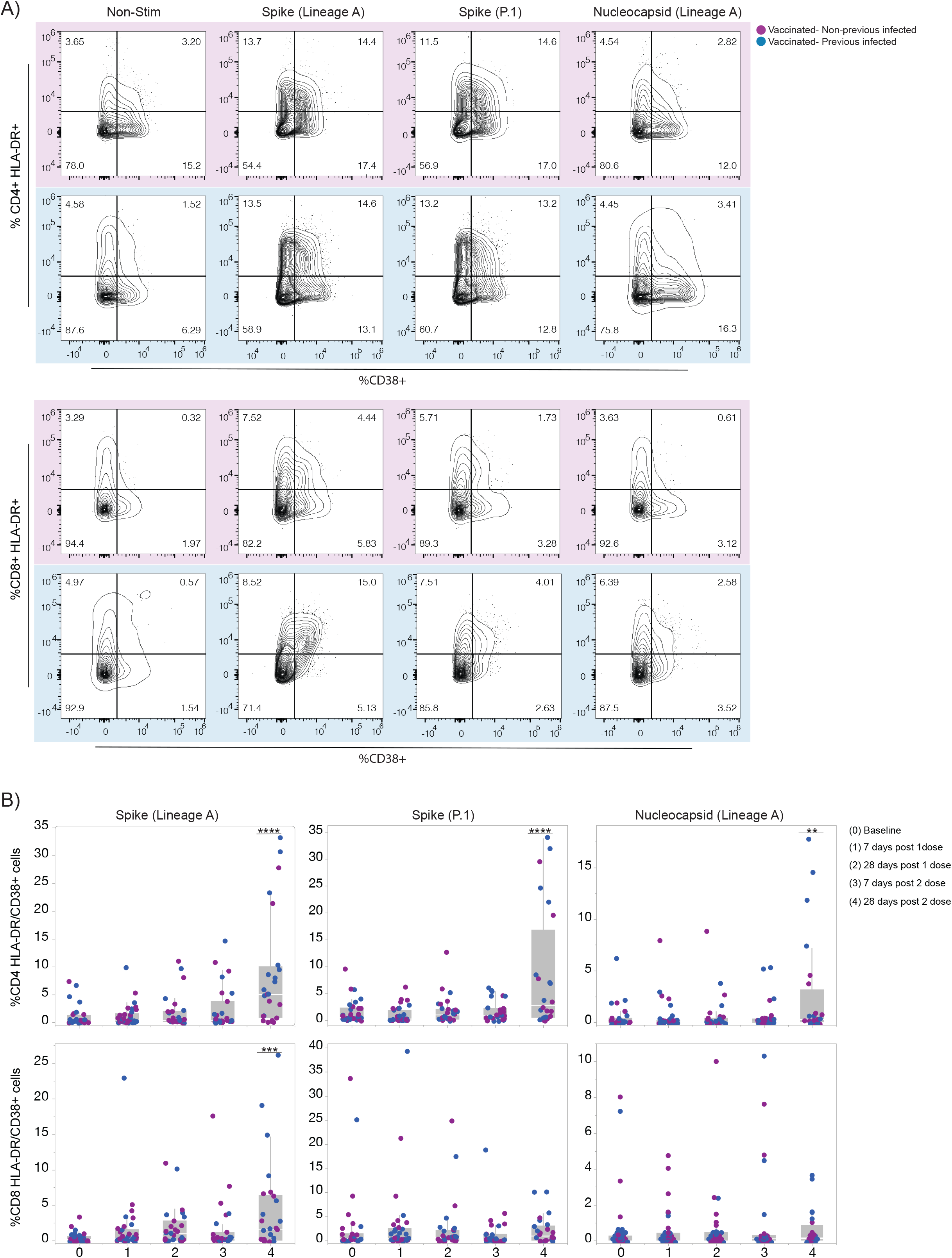
Temporal dynamics of anti-SARS-CoV-2 T cell immunity in vaccinated participants. **a, b**, SARS-CoV-2 S-reactive CD4+ and CD8+T cells after in vitro stimulation with SARS-CoV-2 S-I and S-II peptide pools and Nucleoprotein peptides pool. **a**, Representative dot plots from four vaccinated individuals, 28days post 2 vaccination dose, showing the percentage of double-positive cells expressing HLA-DR and CD38 out of CD4+T cells (top) and CD8+T cells (bottom). Individuals previously infected to SARS-CoV2 or uninfected are indicated by blue or purple shades, respectively. **b**, Percentage of double-positive cells, S-reactive and N-reactive out of CD4+T cells (top) and CD8+T cells (bottom) over time post-vaccination. Individuals previously infected to SARS-CoV2 or uninfected are indicated by blue or purple dots, respectively. Each dot represents a single individual. Significance was assessed by One-way ANOVA corrected for multiple comparisons using Dunnett’s method. Vaccination time points were compared with baseline. Stimulation values were subtracted from the respective non-stimulation condition. Boxes represent variables’ distribution with quartiles and outliers. Horizontal bars, mean values. TP, vaccination time point (TP0, n = 30; TP1, n=34; TP2, n =27; TP3, n = 27; TP4, n=24). Non-Stim, non-stimulated PBMCs. Nucleocapsid, PBMCs stimulated with SARS-CoV-2 nucleocapsid (N) protein pool derived from the ancestral lineage A virus, WA1, USA. Spike, PBMCs stimulated with SARS-CoV-2 spike (S) protein pool derived from the ancestral strain lineage A, WA1, USA. Spike (P.1), PBMCs stimulated with SARS-CoV-2 Spike (S) protein pool derived from the P.1 variant. ****p < .0001***p < .001 **p < .01*p < .05.

To investigate potential differences in NAb escape between the SARS-CoV-2 variants, we analyzed the neutralization capacity of plasma samples from vaccinated individuals against a panel of 18 genetically distinct and authentic SARS-CoV-2 isolates. Among the isolates, 16 were from our Connecticut SARS-CoV-2 genomic surveillance program representing variants from the same geographical region as our HCW cohort^18^. Our variant panel includes representatives of all lineages currently classified as variants of concern (B.1.1.7, B.1.351, P.1, and B.1.617.2) as well as lineages classified as variants of interest (B.1.427, B.1.429, B.1.525, B.1.526, and B.1.617.1)^19^. In addition, we selected lineages with key S gene mutations (B.1.517 with N501T, and B.1 and R.1 with E484K)^20^, and included lineage A as a comparison (Figure 3a). To help deconvolute the effects of individual mutations, we included 4 different B.1.526 isolates (labeled as B.1.526^a-d^) that represent different phylogenetic clades and key S gene mutations (L452R, S477N, and E484K; <u>Extended Data Figure 3b</u>), two different B.1.1.7 isolates with (B.1.1.7^b^) and without E484K (B.1.1.7^a^, most common), and two B.1.351 isolates with (B.1.351^b^) and without L18F (B.1.351^a^, most common; Figure 3a & <u>Extended Data Table 2</u>). Except lineage A, all isolates (lineages B, P, and R) have the S gene D614G mutation located in the receptor-binding motif, which has been reported not to impact vaccine-elicited neutralization^21^. For each isolate we highlight additional key amino acid differences concentrated in the subunit 1, including RBD and additional antigenic sites of the S protein (Figure 3a): L18F and ΔH69/V70 (deletions) located in the amino-terminal domain (NTD); K417N and L452R located in the epitopes of the RBD; S477N, T478K, E484K/Q, and N501T/Y located in the RBD-ACE2 interface; and P681H/R located in the furin cleavage site. A full list of amino acid substitutions and deletions from all genes is provided in <u>Extended Data Table 2</u>. We used a PRNT50 assay to determine the neutralization titers of plasma collected from 32 HCWs 28 days after the second vaccination dose to each isolate.

**Figure 3.**
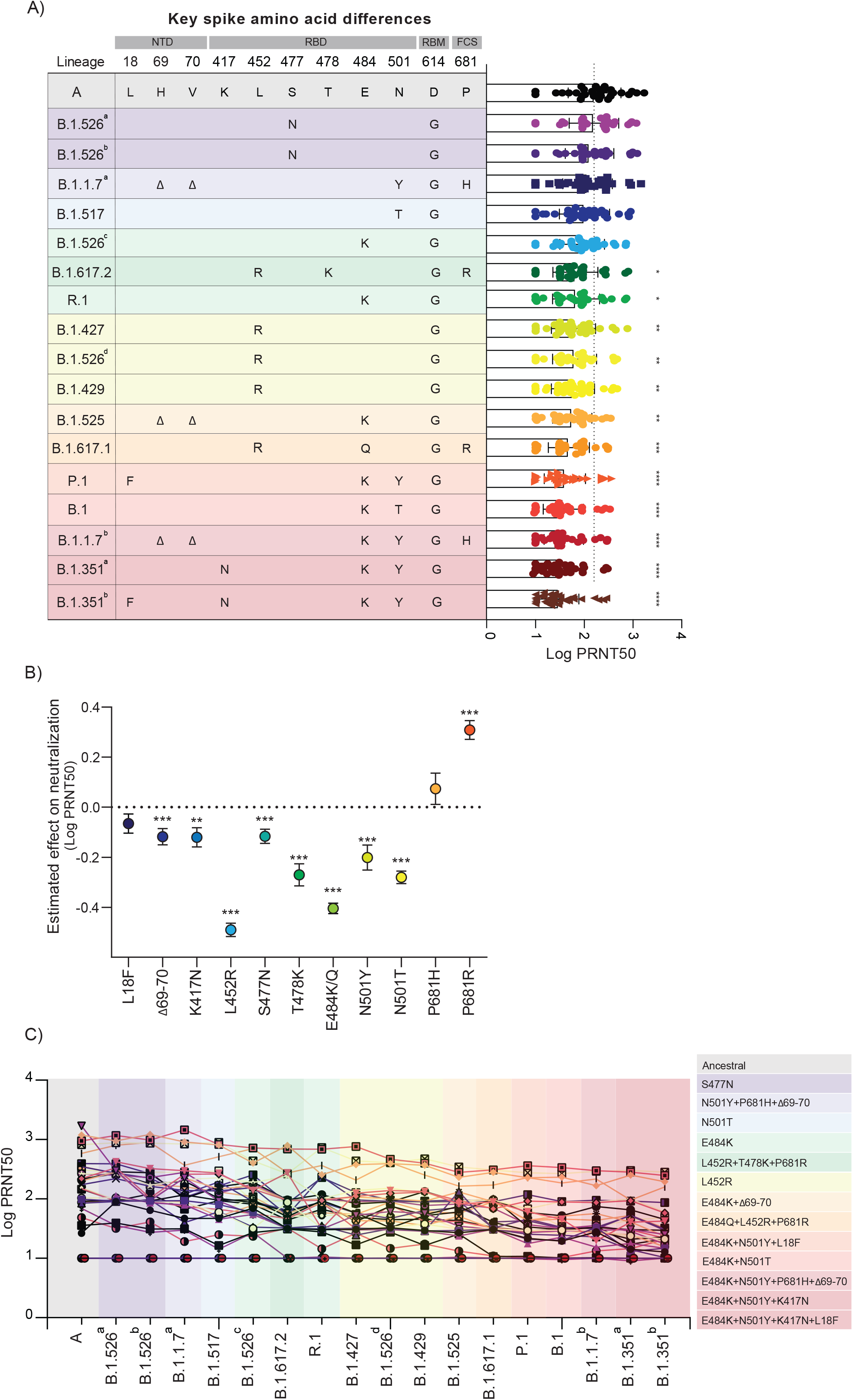
Impact of SARS-CoV-2 VOC on neutralization capacity of vaccinated participants. **a**, Plasma neutralization titers against ancestral lineage A virus, (WA1, USA) and locally circulating variants of concern or interest, and other lineages. Sixteen SARS-Cov2 variants were isolated from nasopharyngeal swabs of infected individuals and an additional B.1.351 isolate was obtained from BEI. Neutralization capacity was accessed using plasma samples from vaccinated participants, 28 days post SARS-CoV2 second vaccination dose at the experimental sixfold serial dilutions (from 1:3 to 1:2430). **a**, Overview of key spike mutations within the distinct lineages and plasma neutralization titers (PRNT50). Spike mutations are arranged across columns and each row represents a SARS-CoV-2 lineage. Significance was assessed by One-way ANOVA corrected for multiple comparisons using Dunnett’s method. Neutralization capacity to the variants was compared to neutralization capacity against the ancestral strain. Boxes represent mean values ± standard deviations. Dotted line indicates the mean value of PRNT50 to ancestral strain. **b**, Estimated effect of individual mutations on plasma neutralization titers. Neutralization estimates (Log PRNT50) and significance were tested with a linear mixed model with subject-level random effects. Dots represent model estimates and error bars show the standard error. ****p < .0001***p < .001 **p < .01*p < .05. **c**, Individual trajectories of plasma neutralization titers (PRNT50). Each line represents a single individual. n=32. Each dot represents a single individual. Dotted line indicates the mean value of PRNT50 to ancestral strain of previously infected individuals, prior to vaccination (baseline). Variants were grouped giving specific-spike mutations and are coloured accordingly. ****p < .0001 ***p < .001 **p < .01*p < .05. NTD, amino-terminal domain. RBD, receptor-binding domain. RBM, receptor-binding motif. FCS, furin cleavage site.

When comparing vaccine-induced neutralization against the different isolates in comparison to the lineage A virus isolate, we observed significantly reduced PRNT50 titers for 12 out of the 17 isolates, and the rank order of reduced neutralization mostly clustered by key S gene amino acid differences (Figure 3a). Virus isolates with both the E484K and N501Y (or N501T) mutations (B.1.351^b^, B.1.351^a^, B.1.1.7^b^, B.1, and P.1) reduced neutralization the most (4.6-6.0 fold decrease in PRNT50 titers). Virus isolates with the L452R mutation (B.1.617.1, B.1.429, B.1.526^b^, B.1.427, and B.1.617.2) were in the next grouping of decreased neutralization (2.5-4.1 fold decrease), which partially overlapped with isolates with E484K but without N501Y/T (B.1.525, R.1, and B.1.526^c^; 2.0-3.8 fold decrease). These data suggest that S gene mutations L452R, E484K, and N501Y have the greatest individual effects on decreasing neutralization. To further assess this possibility, we constructed a linear mixed model with subject-level random effects to account for the differences in neutralization outcome (log transformed PRNT50 titers) by each individual mutation as compared to lineage A (with no mutation; Figure 3b). From our model, we estimated that 8 of the 11 key S gene mutations that we investigated had significant negative effects on neutralization, and that L452R (2.8 fold decrease in PRNT50 titers [mean]; *p*< 2e-16) and E484K/Q (2.0 fold decrease; *p*< 2e-16) had the greatest individual effects. As combinations of mutations can alter effects differently than the added value of each individually (*i*.*e*. epistatic interactions), we also created a second linear mixed model that controlled for all of the individual mutations in the first model as well as three common combinations of key S gene mutations found in our isolates: ΔH69/V70 and E484K, L452R and P681R, and E484K and N501Y. These combinations of mutations allowed us to assess if the contribution of mutations together is synergistic, antagonistic, or neither. Our model suggests that the ΔH69/V70 and E484K combination was synergistic (*i*.*e*. decreased neutralization more than the added effects of each; β=-0.182; *p*=0.005), L452R and P681R was antagonistic (*i*.*e*. decreased neutralization less than the added effects of each; β=0.228; *p*=0.003), and E484K and N501Y was neither (*i*.*e*. neutralization likely the sum of the individual effects of each; β=0.060; *p*=0.248). Thus, from our large panel of virus isolates, we find that virus genotype plays an important role in vaccine-induced neutralization, with L452R likely having the largest individual impact, but the added effects of E484K and N501Y make viruses with this combination perhaps the most concerning for vaccines.

Although virus-specific factors may play a significant role in neutralization, differences in neutralization activity between individual vaccinated HCWs were much larger (up to ∼2 log PRNT50 titers) than differences among virus isolates (mostly <1 log; Figure 3a). By tracking PRNT50 titers from each HCW, we found the vaccinated individuals with high neutralization activity for the lineage A virus isolate are typically on the higher end of neutralization for all variants (Figure 3c). Moreover, two vaccinated HCWs did not develop neutralizing antibodies against any of the virus isolates, including lineage A (Figure 3c), despite the production of virus-specific antibodies (Figure 1a, b).

To further understand the underlying factors that determine levels of neutralization activity, we separated individuals by their SARS-CoV-2 previous infection status (*i*.*e*. previously infected vs uninfected) and determined their neutralization titers to our panel of SARS-CoV-2 isolates. While the rank order of virus isolates impacting neutralization remained mostly the same, we found that the plasma from previously infected vaccinated individuals generally had higher PRNT50 titers against the panel of SARS-CoV-2 isolates than uninfected vaccinated individuals (Figure 4a, b). With the exception of virus isolates from lineages A, B.1.526^a-c^, and R.1, which affected neutralization the least, all other assayed isolates had a significantly higher NAb response in previously infected vaccinated individuals (Figure 4b); only virus isolates with the E484K and N501Y/T mutations still significantly reduced neutralization (Figure 4a). For example, the lineage B.1.351^b^ isolate (E484K and N501Y) decreased neutralization titers by 13.2 fold (compared to lineage A) in uninfected and by 3.7 fold in previously infected vaccinated individuals, whereas B.1.617.2 (L452R) went from 6.9 to 1.5 and B.1.1.7^a^ (N501Y) went from 3.4 to 0.8 fold decrease in uninfected to infected, respectively (Figure 4a). Thus, our data suggests that plasma neutralization activity against SARS-CoV-2 variants is improved in vaccinated individuals previously infected with the virus. These results suggest that future vaccine boosters (third dose) may help to overcome the reduction in neutralization capacity observed for the variants, in particular those of the genotypes with the L452R (*e*.*g*., B.1.617.2) or the E484K and N501Y combination (*e*.*g*., P.1 and B.1.351).

**Figure 4.**
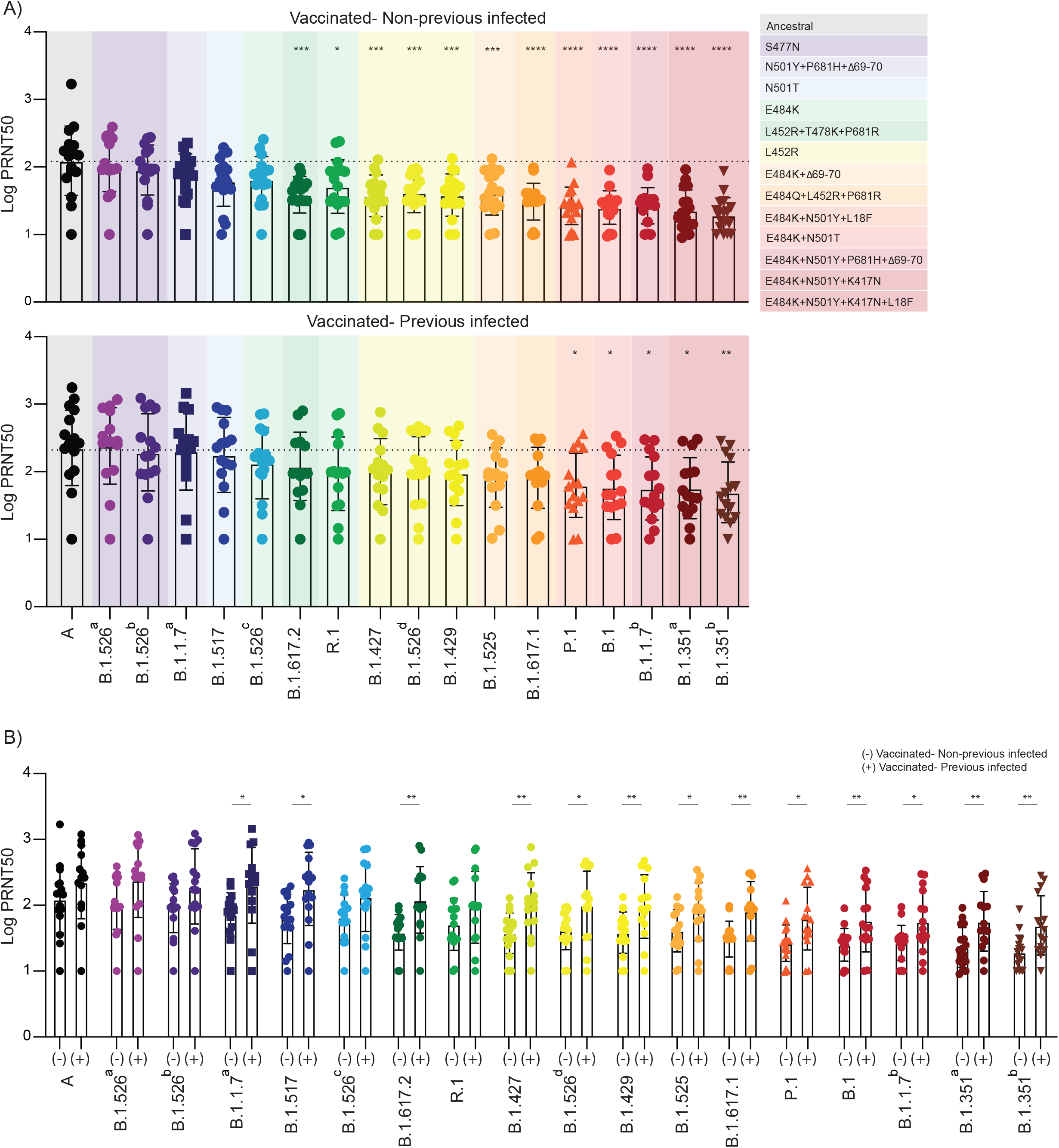
Neutralizing activity comparison in vaccinated healthcare workers previously infected or not to SARS-CoV-2. **a-b**, Plasma neutralization titers against ancestral lineage A virus, (WA1, USA) and locally circulating variants of concern or interest, and other lineages. Sixteen SARS-Cov2 variants were isolated from nasopharyngeal swabs of infected individuals and an additional B.1.351 isolate was obtained from BEI. Neutralization capacity was accessed using plasma samples from vaccinated participants, 28 days post SARS-CoV2 second vaccination dose at the experimental sixfold serial dilutions (from 1:3 to 1:2430). **a, b**, Neutralization capacity between vaccinated participants previously infected or not to SARS-CoV-2. **a**, Neutralization titer among vaccinated individuals. Significance was assessed by One-way ANOVA corrected for multiple comparisons using Dunnett’s method. Neutralization capacity to the variants was compared to neutralization capacity against the ancestral strain. Boxes represent mean values ± standard deviations. Dotted line indicates the mean value of PRNT50 to ancestral strain. Variants were grouped giving specific-spike mutations and are coloured accordingly. ****p < .0001 ***p < .001 **p < .01*p < .05. **b**, Neutralization titer comparison among vaccinated participants previously infected or not to SARS-CoV-2. Significance was accessed using unpaired t-test. Boxes represent mean values ± standard deviations. (-) Vaccinated-uninfected, n=17; (+) Vaccinated-Previously infected, n=15. Each dot represents a single individual **p < .01*p < .05.

## Discussion

Human NAbs against SARS-CoV-2 can be categorized as belonging to four classes on the basis of their target regions on the RBD. Although the RBD is immunodominant, there is evidence for a substantial role of other spike regions in antigenicity, most notably the NTD supersite^22–24^. These antibodies target epitopes that are closely associated with NTD and RBD residues L18 and ΔH69/V70, and K417, L452, S477, T478, E484 and N501. Previous studies using pseudovirus constructs reported a significant impact of single S amino acid substitutions, including S477N, E484K, and N501Y, located at the RBD-ACE2 interface, in the neutralization activity of plasma from vaccinated individuals^1–5^.

Using a large panel of genetically diverse authentic SARS-CoV-2 isolates, we found that decreases in mRNA vaccine-induced neutralization capacity could be generally categorized into three tiers based on the presence of L452R, E484K, and/or N501Y/T. Tier 1, which we found to decrease neutralization titers by >10 fold in previously uninfected vaccinated individuals, includes lineages with both the E484K and N501Y/T mutations such as B.1.351 (Beta) and P.1 (Gamma), further supporting their importance in regards to vaccines. Interestingly, we also found that a generic lineage B.1 isolate with E484K and N501T, and a rare B.1.1.7 (Alpha) isolate with E484K (also with the common N501Y mutation) have similar impacts on neutralization as B.1.351 and P.1. While the combinations of mutations in the B.1 and B.1.1.7 with E484K isolates likely do not increase transmissibility, the additive effects of these two mutations supports that surveillance programs should track all viruses with E484K and N501Y/T in addition to variants of concern/interest.

Tier 2 consists of SARS-CoV-2 lineages with the L452R mutation, and tier 3 consists of lineages with E484K but without N501Y/T. Notably, lineages with the L452R mutation include B.1.617.2 (Delta). We found that B.1.617.2 had a comparatively modest impact on neutralization titers: 6.9 fold decrease in uninfected and 1.5 fold decrease in previously infected vaccinated individuals (2.5 fold decrease combined) as compared to lineage A. From these data, we expect that most fully vaccinated individuals will be protected against B.1.617.2, and that the rise in vaccine-breakthroughs associated with this variant are more likely associated with its high transmissibility^25–28^. Our inclusion of 4 different versions of B.1.526 (Iota, first detected in New York) shows hierarchy of viruses with L452R, S477N, or E484K with regards to vaccine neutralization. While some have speculated that B.1.526 with E484K may have the greatest public health implications^29,30^, we found that B.1.526 with L452R decreases neutralization more than the other versions. Overall, we found that L452R has the greatest single effect on neutralization across several lineages, and viruses without L452R or E484K have insignificant effects on neutralization (this includes B.1.526 with S477N).

The discrepancies of our results compared to other studies may point to the importance of using fully intact authentic virus for neutralization assays to detect effects of epistasis among virus mutations on neutralization assays. Nevertheless, it remains possible that additional factors also contribute to some of the discrepancies between our observations and those of previous studies, including the composition of our cohorts, predominantly young Caucasian women. Discrepancies between cohorts could also account for subtle differences in T cell responses observed in our study versus the ones recently reported by Alter et al., and Tarke et al. While we observed decreased cross-reactivity of S-reactive CD8^+^ T cells against P.1 S peptides, the above studies found T cell responses are largely preserved against variants^16,17^. In addition to cohort composition, we used overlapping peptide pools in our assays, hence, we cannot rule out the possibility that some mutations affect antigen processing prior to presentation. Furthermore, Alter et al. used an AdV1.26 vaccine^16^, while ours and Tarke et al. used mRNA vaccines^17^. A potential limitation in our study is that, even with our 7-day stimulation prior to analysis, it remains possible that our T cells assay failed to detect underrepresented T cell clones impacted by variant sequences when sampled in the presence of the majority of conserved peptides. Overall, our data point to a necessity of active monitoring of T cell reactivity in the context of SARS-CoV-2 evolution.

The magnitude of the antibody titers in COVID-19 patients following natural infection has been directly correlated with length of infection and severity^31^. Here, we found that previously infected vaccinated individuals display an increased resilience in antibody responses against both “single” and combination of substitutions in the RBD region, which otherwise severely decreased neutralization activity of uninfected vaccinated individuals. Our observations of the impact of pre-existing immunity in vaccinated individuals on their ability to neutralize variants could be explained by the time window between the initial exposure (infection) and vaccination. Still, our observations provide an important rationale for worldwide efforts in characterizing the contribution of pre-existing SARS-CoV-2 immunity to the outcome of various vaccination strategies. Along with recently introduced serological tests^32^, such studies could inform evidence-based risk evaluation, patient monitoring, adaptation of containment methods and vaccine development and deployment. Finally, these findings suggest that an additional third vaccination dose may be beneficial to confer higher protection against SARS-CoV-2 lineages such as B.1.351 and P.1.

## METHODS

### Ethics statement

This study was approved by Yale Human Research Protection Program Institutional Review Board (IRB Protocol ID 2000028924). Informed consent was obtained from all enrolled vaccinated HCWs. The Institutional Review Board from the Yale University Human Research Protection Program determined that the RT-qPCR testing and sequencing of de-identified remnant COVID-19 clinical samples conducted in this study is not research involving human subjects (IRB Protocol ID: 2000028599).

### Healthcare workers volunteers

Forty health care workers volunteers from the YNHH were enrolled and included in this study. The volunteers received the mRNA vaccine (Moderna or Pfizer) between November 2020 and January 2021. Vaccinated donors were stratified in two major groups, previously infected with SARS-CoV2 (recovered) on uninfected (naive), confirmed by RT-qPCR and serology. None of the participants experienced serious adverse effects after vaccination. HCWs were followed serially post-vaccination. Plasma and PBMCs samples were collected at baseline (previous to vaccination), 7- and 28- post first vaccination dose, and 7-, 28- and 70-days post second vaccination dose. Demographic information was aggregated through a systematic review of the EHR and was used to construct <u>Extended Data Table 1</u>. The clinical data were collected using EPIC EHR May 2020 and REDCap 9.3.6 software. Blood acquisition was performed and recorded by a separate team. Vaccinated HCW’s clinical information and time points of collection information was not available until after processing and analyzing raw data by flow cytometry and ELISA. ELISA, neutralizations, and flow cytometry analyses were blinded.

### Isolation of plasma and PBMCs

Whole blood was collected in heparinized CPT blood vacutainers (BD; # BDAM362780) and kept on gentle agitation until processing. All blood was processed on the day of collection in a single step standardised method. Plasma samples were collected after centrifugation of whole blood at 600 g for 20 min at room temperature (RT) without brake. The undiluted plasma was transferred to 15-ml polypropylene conical tubes, and aliquoted and stored at −80 °C for subsequent analysis. The PBMC layer was isolated according to the manufacturer’s instructions. Cells were washed twice with PBS before counting. Pelleted cells were briefly treated with ACK lysis buffer for 2 min and then counted. Percentage viability was estimated using standard Trypan blue staining and an automated cell counter (Thermo-Fisher, #AMQAX1000). PBMCs were stored at -80 °C for subsequent analysis.

### SARS-CoV-2 specific-antibody measurements

ELISAs were performed as previously described^40^. In short, Triton X-100 and RNase A were added to serum samples at final concentrations of 0.5% and 0.5mg/ml respectively and incubated at room temperature (RT) for 30 minutes before use, to reduce risk from any potential virus in serum. 96-well MaxiSorp plates (Thermo Scientific #442404) were coated with 50 μl/well of recombinant SARS Cov-2 STotal (ACROBiosystems #SPN-C52H9-100ug), S1 (ACROBiosystems #S1N-C52H3-100ug), RBD (ACROBiosystems #SPD-S52H6-100ug) and Nucleocapsid protein (ACROBiosystems #S1N-C52H3-100ug) at a concentration of 2 μg/ml in PBS and were incubated overnight at 4 °C. The coating buffer was removed, and plates were incubated for 1 h at RT with 200 μl of blocking solution (PBS with 0.1% Tween- 20, 3% milk powder). Plasma was diluted serially 1:100, 1:200, 1:400 and 1:800 in dilution solution (PBS with 0.1% Tween-20, 1% milk powder) and 100 μl of diluted serum was added for two hours at RT. Human Anti-Spike and anti-nucleocapsid antibodies were serially diluted to generate a standard curve. Plates were washed three times with PBS-T (PBS with 0.1% Tween-20) and 50 μl of HRP anti-Human IgG Antibody (GenScript #A00166, 1:5,000) diluted in dilution solution added to each well. After 1 h of incubation at RT, plates were washed six times with PBS-T. Plates were developed with 100 μl of TMB Substrate Reagent Set (BD Biosciences #555214) and the reaction was stopped after 5 min by the addition of 2 N sulfuric acid. Plates were then read at a wavelength of 450 nm and 570nm.

### T cells stimulation

For the *in vitro* stimulation, PBMCs were stimulated with HLA class I and HLA-DR peptide pools at the concentration of 1-10 μg/ml per peptide and cultured for 7 days. On day 0, PBMCs were thawed, counted, and plated in a total of 5–8×10^5^ cells per well in 200ul of RPMI 1640 medium (Gibco) supplemented with 1% sodium pyruvate (NEAA), 100 U/ml penicillin /streptomycin (Biochrom) and 10% fetal bovine serum (FBS) at 37°C and 5% CO2. On day 1, cells were washed and the stimulation was performed with: PepMix SARS-CoV-2 spike glycoprotein pool 1 and pool 2 (GenScript), PepMix P.1 SARS-CoV-2 spike glycoprotein pool 1 and pool 2 (JPT) and PepMix SARS-CoV-2 Nucleocapsid protein (JPT). Stimulation controls were performed with PBS (unstimulated). Peptide pools were used at 1 μg/ml per peptide. Incubation was performed at 37□°C, 5% CO2 for 6 days. On day 6, cells were restimulated with 10ug/ml per peptide and subsequently incubated for 12h, being the last 6 h in the presence of 10 μg/ml brefeldin A (Sigma-Aldrich). Following this incubation, cells were washed with PBS 2 mM EDTA and prepared for analysis by flow cytometry.

### Flow cytometry

Antibody clones and vendors were as follows: BB515 anti-hHLA-DR (G46-6) (1:400) (BD Biosciences), BV605 anti-hCD3 (UCHT1) (1:300) (BioLegend), BV785 anti-hCD19 (SJ25C1) (1:300) (BD Biosciences), BV785 anti-hCD4 (SK3) (1:200) (BioLegend), APCFire750 or BV711 anti-hCD8 (SK1) (1:200) (BioLegend), AlexaFluor 700 anti-hCD45RA (HI100) (1:200) (BD Biosciences), PE anti-hPD1 (EH12.2H7) (1:200) (BioLegend), APC or PE-CF594 anti-hTIM3 (F38-2E2) (1:50) (BioLegend), BV711 anti-hCD38 (HIT2) (1:200) (BioLegend), BB700 anti-hCXCR5 (RF8B2) (1:50) (BD Biosciences), PE-CF594 anti-hCD25 (BC96) (1:200) (BD Biosciences), AlexaFluor 700 anti-hTNFa (MAb11) (1:100) (BioLegend), PE or APC/Fire750 anti-hIFNy (4S.B3) (1:60) (BioLegend), FITC anti-hGranzymeB (GB11) (1:200) (BioLegend), BV785 anti-hCD19 (SJ25C1) (1:300) (BioLegend), BV421 anti-hCD138 (MI15) (1:300) (BioLegend), AlexaFluor700 anti-hCD20 (2H7) (1:200) (BioLegend), AlexaFluor 647 anti-hCD27 (M-T271) (1:350) (BioLegend), PE/Dazzle594 anti-hIgD (IA6-2) (1:400) (BioLegend), Percp/Cy5.5 anti-hCD137 (4B4-1) (1:150) (BioLegend) and PE anti-CD69 (FN-50) (1:200) (BioLegend), APC anti-hCD40L (24-31) (1:100) (BioLegend). In brief, freshly isolated PBMCs were plated at 1–2 × 10^6^ cells per well in a 96-well U-bottom plate. Cells were resuspended in Live/Dead Fixable Aqua (ThermoFisher) for 20 min at 4°C. Following a wash, cells were blocked with Human TruStan FcX (BioLegend) for 10 min at RT. Cocktails of desired staining antibodies were added directly to this mixture for 30 min at RT. For secondary stains, cells were first washed and supernatant aspirated; then to each cell pellet a cocktail of secondary markers was added for 30 min at 4 °C. Prior to analysis, cells were washed and resuspended in 100 μl 4% PFA for 30 min at 4 °C. Following this incubation, cells were washed and prepared for analysis on an Attune NXT (ThermoFisher). Data were analysed using FlowJo software version 10.6 software (Tree Star). The specific sets of markers used to identify each subset of cells are summarized in <u>Extended Data Figure 4.</u>

### Cell lines and virus

TMPRSS2-VeroE6 kidney epithelial cells were cultured in Dulbecco’s Modified Eagle Medium (DMEM) supplemented with 1% sodium pyruvate (NEAA) and 5% fetal bovine serum (FBS) at 37°C and 5% CO2. The cell line was obtained from the ATCC and has been tested negative for contamination with mycoplasma. SARS-CoV-2 lineage A(USA-WA1/2020), was obtained from BEI Resources (#NR-52281) and was amplified in TMPRSS2-VeroE6. Cells were infected at a MOI 0.01 for three days to generate a working stock and after incubation the supernatant was clarified by centrifugation (450 g × 5 min), and filtered through a 0.45-micron filter. The pelleted virus was then resuspended in PBS and aliquoted for storage at −80°C. Viral titers were measured by standard plaque assay using TMPRSS2-VeroE6. Briefly, 300 µl of serial fold virus dilutions were used to infect Vero E6 cells in MEM supplemented NaHCO3, 4% FBS 0.6% Avicel RC-581. Plaques were resolved at 48h post-infection by fixing in 10% formaldehyde for 1h followed by 0.5% crystal violet in 20% ethanol staining. Plates were rinsed in water to plaques enumeration. All experiments were performed in a biosafety level 3 laboratory with approval from the Yale Environmental Health and Safety office.

### SARS-CoV-2 Variant sequencing and isolation

SARS-CoV-2 samples were sequenced as part of the Yale Genomic Surveillance Initiative’s weekly surveillance program in Connecticut, United States^33^. Lineages were sequenced and isolated as described previously^34^. In brief, nucleic acid was extracted from de-identified remnant nasopharyngeal swabs and tested with our multiplexed RT-qPCR variant assay to select samples with a N1 cycle threshold (Ct) value of 35 or lower for sequencing^35,36^. Libraries were prepared with a slightly adjusted version of the Illumina COVIDSeq Test RUO version. The Yale Center for Genome Analysis sequenced pooled libraries of up to 96 samples on the Illumina NovaSeq (paired-end 150). Data was analyzed and consensus genomes were generated using iVar (version 1.3.1)^37^. Variants of interest and concern, lineages with mutations of concern (E484K), as well as other lineages as controls were selected for virus isolation. In total, 16 viruses were isolated belonging to 12 lineages <u>(Extended Data Figure 3; Extended Data Table 2)</u>. In addition, ancestral lineage A virus and lineage B.1.351 virus were obtained from BEI.

Samples selected for virus isolation were diluted 1:10 in Dulbecco’s Modified Eagle Medium (DMEM) and then filtered through a 45 µM filter. The samples were ten-fold serially diluted from 1:50 to 1:19,531,250. The dilution was subsequently incubated with TMPRSS2-Vero E6 in a 96 well plate and adsorbed for 1 hour at 37°C. After adsorption, replacement medium was added, and cells were incubated at 37°C for up to 5 days. Supernatants from cell cultures with cytopathic effect (CPE) were collected, frozen, thawed and subjected to RT-qPCR. Fresh cultures were inoculated with the lysates as described above for viral expansion. Viral infection was subsequently confirmed through reduction of Ct values in the cell cultures with the multiplex variant qPCR assay. Expanded viruses were re-sequenced following the same method as described above and genome sequences were uploaded to GenBank (<u>Supplementary Data Table 2</u>), and the aligned consensus genomes are available on GitHub (https://github.com/grubaughlab/paper_2021_Nab-variants). Nextclade v1.5.0 (https://clades.nextstrain.org/) was used to generate a phylogenetic tree <u>(Extended Data Figure 3),</u> and to compile a list of amino acid changes in the virus isolates as compared to the Wuhan-Hu-1 reference strain (Extended Data Table 2). Key spike amino acid differences were identified based on the outbreak.info mutation tracker^20^.

### Neutralization assay

Vaccinated HCWs sera were isolated as described before and then heat treated for 30 min at 56°C. Sixfold serially diluted plasma, from 1:3 to 1:2430 were incubated with SARS-CoV-2 variants, for 1 h at 37□°C. The mixture was subsequently incubated with TMPRSS2-VeroE6 in a 12-well plate for 1h, for adsorption. Then, cells were overlayed with MEM supplemented NaHCO_3_, 4% FBS 0.6% Avicel mixture. Plaques were resolved at 40 h post infection by fixing in 10% formaldehyde for 1 h followed by staining in 0.5% crystal violet. All experiments were performed in parallel with baseline controls sera, in an established viral concentration to generate 60-120 plaques/well.

### Statistical analysis

All analyses of patient samples were conducted using GraphPad Prism 8.4.3, JMP 15, and R 3.4.3. Multiple group comparisons were analyzed by running parametric (ANOVA) statistical tests. Multiple comparisons were corrected using Tukey’s and Dunnett’s test as indicated in figure legends. For the comparison between stable groups, two-sided unpaired t-test was used for the comparison. The impact of spike mutations were assessed using a linear mixed model with an outcome of log transformed PRNT50 and random effects accounting for each individual subject. This was done using the “lme4” package in R 4.0.1^38^.

## Data Availability

All the background information for HCWs participants in this study are included in Source Data Figure 1. All the genome information for SARS-CoV-2 variants used in this study are available in Source Data Figure 2, and the aligned consensus genomes are available on GitHub (https://github.com/grubaughlab/paper_2021_Nab-variants). Additional correspondence and requests for materials should be addressed to the corresponding author (A.I).

## Data availability

All the background information for HCWs participants in this study are included in <u>Source Data Figure 1.</u> All the genome information for SARS-CoV-2 variants used in this study are available in <u>Source Data Figure 2</u>, and the aligned consensus genomes are available on GitHub (https://github.com/grubaughlab/paper_2021_Nab-variants). Additional correspondence and requests for materials should be addressed to the corresponding author (A.I).

## Acknowledgements

We thank M. Linehan for technical and logistical assistance, D. Mucida for discussions, and M. Suchard and W. Hanage for statistics advice. This work was supported by the Women’s Health Research at Yale Pilot Project Program (A.I.), Fast Grant from Emergent Ventures at the Mercatus Center (A.I. and N.D.G.), Mathers Foundation, and the Ludwig Family Foundation, the Department of Internal Medicine at the Yale School of Medicine, Yale School of Public Health and the Beatrice Kleinberg Neuwirth Fund. A.I. is an Investigator of the Howard Hughes Medical Institute. C.L. is a Pew Latin American Fellow. C.B.F.V. is supported by NWO Rubicon 019.181EN.004.

## Author contributions

C.L., C.B.F.V., I.Y., S.O., N.D.G., and A.I. conceived the study. C.L., J.S., S.T., S.M. and J.H collected and processed patient PBMC and plasma samples. C.L. and V.M. performed SARS-CoV-2 specific antibody ELISAs. C.L isolated SARS-CoV-2 variants and performed the neutralization assays. C.L, P.L, and J.G performed the flow cytometry and the flow data analyses. I.Y and M.C. collected epidemiological and clinical data. C.B.F.V., M.I.B., J.R.F., and Y.S.G.S.N. performed virus sequencing. M.C.M processed and stored patient specimens. M.C. assisted volunteers’ identification and enrolment. C.L., C.B.F.V., J.E.R., and N.D.G. analyzed the data. C.L., C.B.F.V., N.D.G and A.I. drafted the manuscript. All authors reviewed and approved the manuscript. N.D.G and A.I. secured funds and supervised the project.

## Competing interests

AI served as a consultant for Spring Discovery, Boehringer Ingelheim and Adaptive Biotechnologies. IY reported being a member of the mRNA-1273 Study Group and has received funding to her institution to conduct clinical research from BioFire, MedImmune, Regeneron, PaxVax, Pfizer, GSK, Merck, Novavax, Sanofi-Pasteur, and Micron. NDG is a consultant for Tempus Labs to develop infectious disease diagnostic assays.

All other authors declare no competing interests.

**Extended Data Figure 1** | **Correlation of virus-specific antibodies with age and sex of participants. a**,**b**, Plasma reactivity to S protein and RBD in vaccinated participants measured over time by ELISA. HCW participants received 2 doses of the mRNA vaccines and plasma samples were collected as at the indicated time points (TP). Baseline, previously to vaccination; 1 Time point, 7 days post 1 dose; 2 Time point, 28 days post 1 dose; 3 Time point, 7 days post 2 dose; 4 Time point, 28 days post 2 dose; 5 Time point, 70 days post 2 dose. **a**, Anti-S (left) and Anti-RBD (right) IgG levels stratified by vaccinated participants accordingly to age and sex. Significance was accessed using unpaired t-test. Boxes represent variables’ distribution with quartiles and outliers. Horizontal bars, mean values. **b**,Anti-S, Anti-S1, Anti-RBD and Anti-N IgG comparison in vaccinated participants previously infected or not to SARS-CoV-2. Longitudinal data plotted over time. Significance was accessed using unpaired t-test. Boxes represent mean values ± standard deviations. TP, vaccination time point. Anti-S IgG (TP0, n = 37; TP1, n=35; TP2, n =30; TP3, n = 34; TP4, n=34; TP5, N=28). Anti-S1 IgG (TP0, n = 37; TP1, n=35; TP2, n =30; TP3, n = 34; TP4, n=34; TP5, N=27). Anti-RBD IgG (TP0, n = 37; TP1, n=35; TP2, n =30; TP3, n = 34; TP4, n=34; TP5, N=27). Anti-N IgG (TP0, n = 37; TP1, n=35; TP2, n =30; TP3, n = 34; TP4, n=34; TP5, N=27). S, spike. S1, spike subunit 1. RBD, receptor binding domain. N, nucleocapsid. Each dot represents a single individual. ****p < .0001 ***p < .001 **p < .01*p < .05.

**Extended Data Figure 2** | **Cellular immune profiling post SARS-CoV2 vaccination. a-b**, Immune cell subsets of interest, plotted as a percentage of a parent population over time according to the vaccination time points. HCW participants received 2 doses of the mRNA vaccines and PBMCs samples were collected as at the indicated time points (TP). Baseline, previously to vaccination; 1 Time point, 7 days post 1 dose; 2 Time point, 28 days post 1 dose; 3 Time point, 7 days post 2 dose; 4 Time point, 28 days post 2 dose. Percentage of activated T cell subsets **(a-b)**, B cell subsets **(c)** and Tfh cells **(d)** among vaccinated individuals over time. Individuals previously infected to SARS-CoV2 or uninfected are indicated by blue or purple dots, respectively. Each dot represents a single individual. Significance was assessed by One-way ANOVA corrected for multiple comparisons using Dunnett’s method. Vaccination time points were compared with baseline. Boxes represent variables’ distribution with quartiles and outliers. Horizontal bars, mean values.TP, vaccination time point (TP0, n = 29; TP1, n=33; TP2, n =26; TP3, n = 13; TP4, n=25). ***p < .001 **p < .01*p < .05.

**Extended Data Figure 3** | **Maximum likelihood phylogeny of SARS-CoV-2 genomes of cultured virus isolates. a**, Nextclade (https://clades.nextstrain.org/) was used to generate a phylogenetic tree to show evolutionary relations between the cultured virus isolates used in this study and other publicly available SARS-CoV-2 genomes. Branches are colored by Pango Lineage, and labelled according to the WHO naming scheme. Highlighted are the cultured virus isolates used in this study. **b**, Enlarged section of the phylogenetic tree highlighting spike amino acid changes in the B.1.526 (iota) lineage viruses belonging to different clades.

**Extended Data Figure 4** | **Gating strategies**. Gating strategies are shown for the key cell populations described in Figure 2 and Extended Data Figure 2. **a**, Leukocyte gating strategy to identify lymphocytes. T cell surface staining gating strategy to identify CD4 and CD8 T cells, TCR-activated T cells and follicular T cells. **b**, B cell surface staining gating strategy to identify B cells subsets.

**Extended Data Table 1** | **SARS-CoV-2 Vaccinated Cohort**. Exact counts for each demographic category are displayed in each cell with accompanying standard deviations for each measurement. Percentages of total, where applicable, are provided in parenthesis. In cases where specific demographic information was missing, the total number of patients with complete information used for calculations is provided within the cell.

**Extended Data Table 2** | **Amino acid changes identified in cultured SARS-CoV-2 isolates**. Cultured virus isolates were resequenced and the consensus genomes were compared to the reference genome (Accession MN908947) using Nextclade (https://clades.nextstrain.org/). Letters indicate amino acids, numbers indicate amino acid positions, asterisks indicate stop codon mutations, and dashes indicate deletions.

**Source Data Figure 1** | **Detailed clinical and immunological data for each patient**. Clinical information, demographics and exact counts for immunological data.

