## Supplementary Figure1, Supplementary Figure2,Supplementary Figure3,Supplementary Figure4,Supplementary Table1,Supplementary Table2 for "Impact of circulating SARS-CoV-2 variants on mRNA vaccine-induced immunity in uninfected and previously infected individuals"

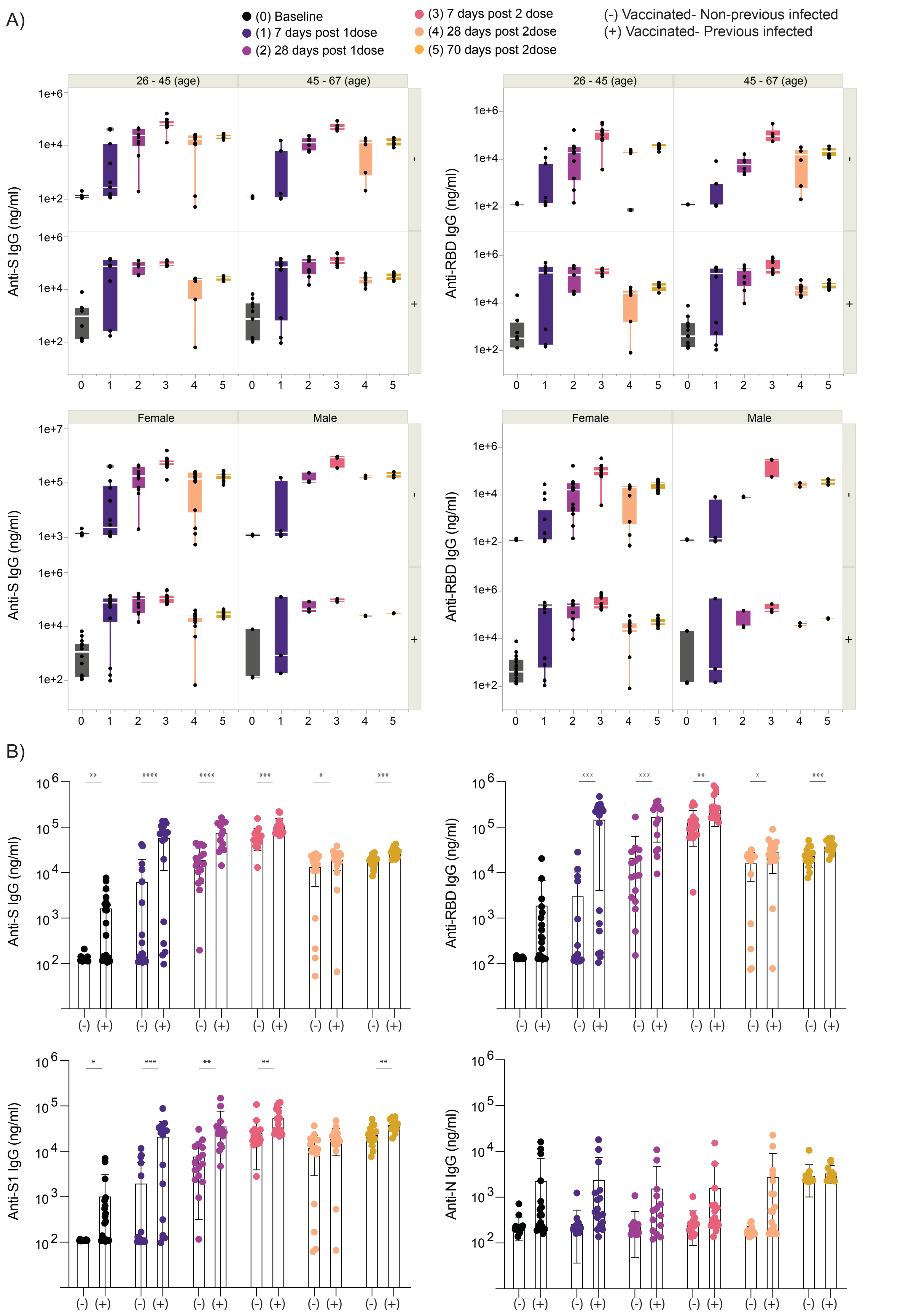

A)
(0) Baseline
(1) 7 days post 1dose
(2) 28 days post 1dose
(3) 7 days post 2 dose
(4) 28 days post 2dose
(5) 70 days post 2dose
(-) Vaccinated- Non-previous infected
(+) Vaccinated- Previous infected
26 - 45 (age)
45 - 67 (age)
Female
Male
Anti-S IgG (ng/ml)
Anti-RBD IgG (ng/ml)
B)
Anti-S IgG (ng/ml)
Anti-RBD IgG (ng/ml)
Anti-S1 IgG (ng/ml)
Anti-N IgG (ng/ml)

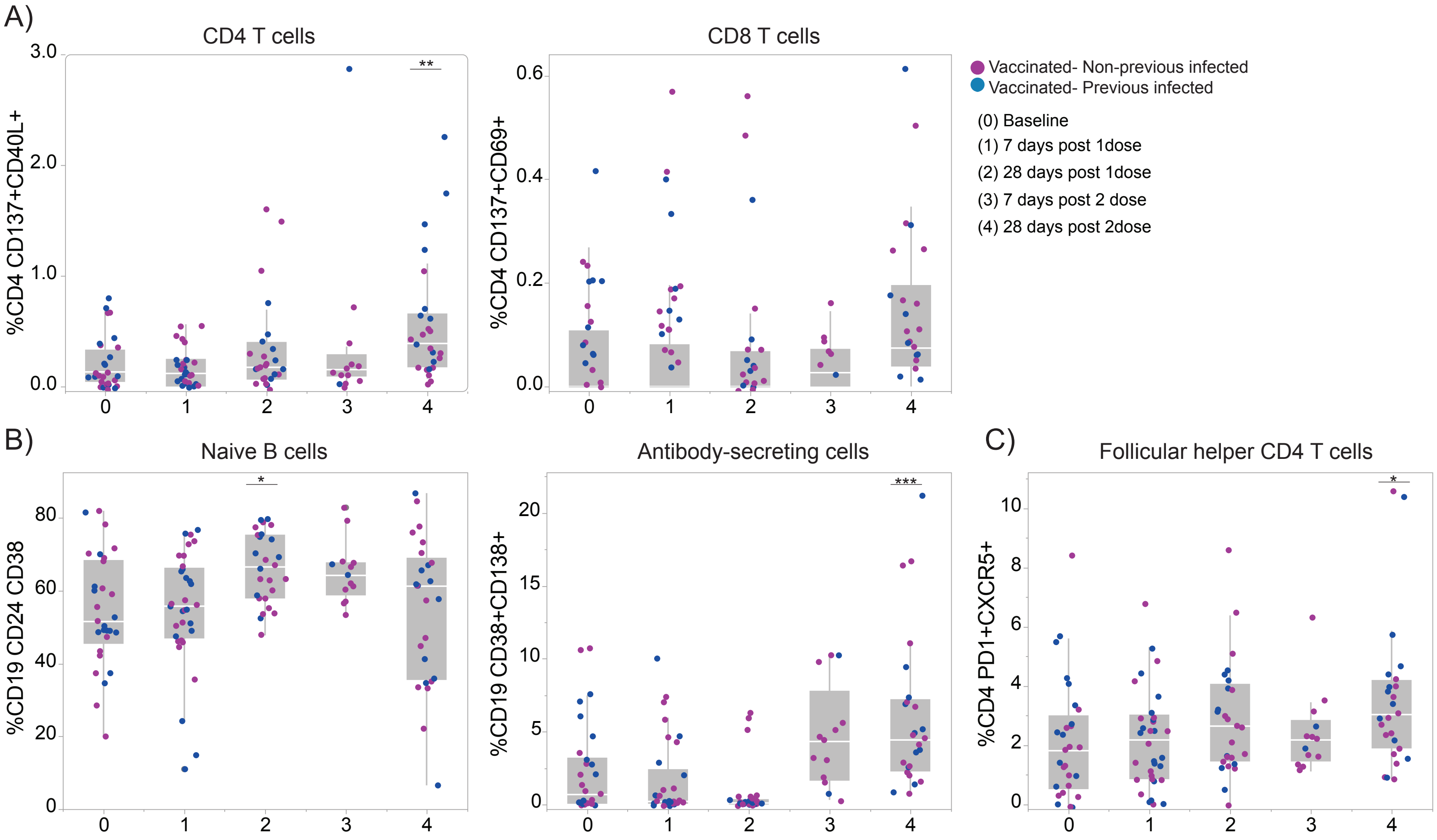
A)
CD4 T cells
%CD4 CD137+CD40L+
3.0
2.0
1.0
0.0
**
0
1
2
3
4
CD8 T cells
%CD4 CD137+CD69+
0.6
0.4
0.2
0.0
Vaccinated- Non-previous infected
Vaccinated- Previous infected
(0) Baseline
(1) 7 days post 1dose
(2) 28 days post 1dose
(3) 7 days post 2 dose
(4) 28 days post 2dose
B)
Naive B cells
%CD19 CD24 CD38
80
60
40
20
0
*
Antibody-secreting cells
%CD19 CD38+CD138+
20
15
10
5
0
***
C)
Follicular helper CD4 T cells
%CD4 PD1+CXCR5+
10
8
6
4
2
0
*

A)

B

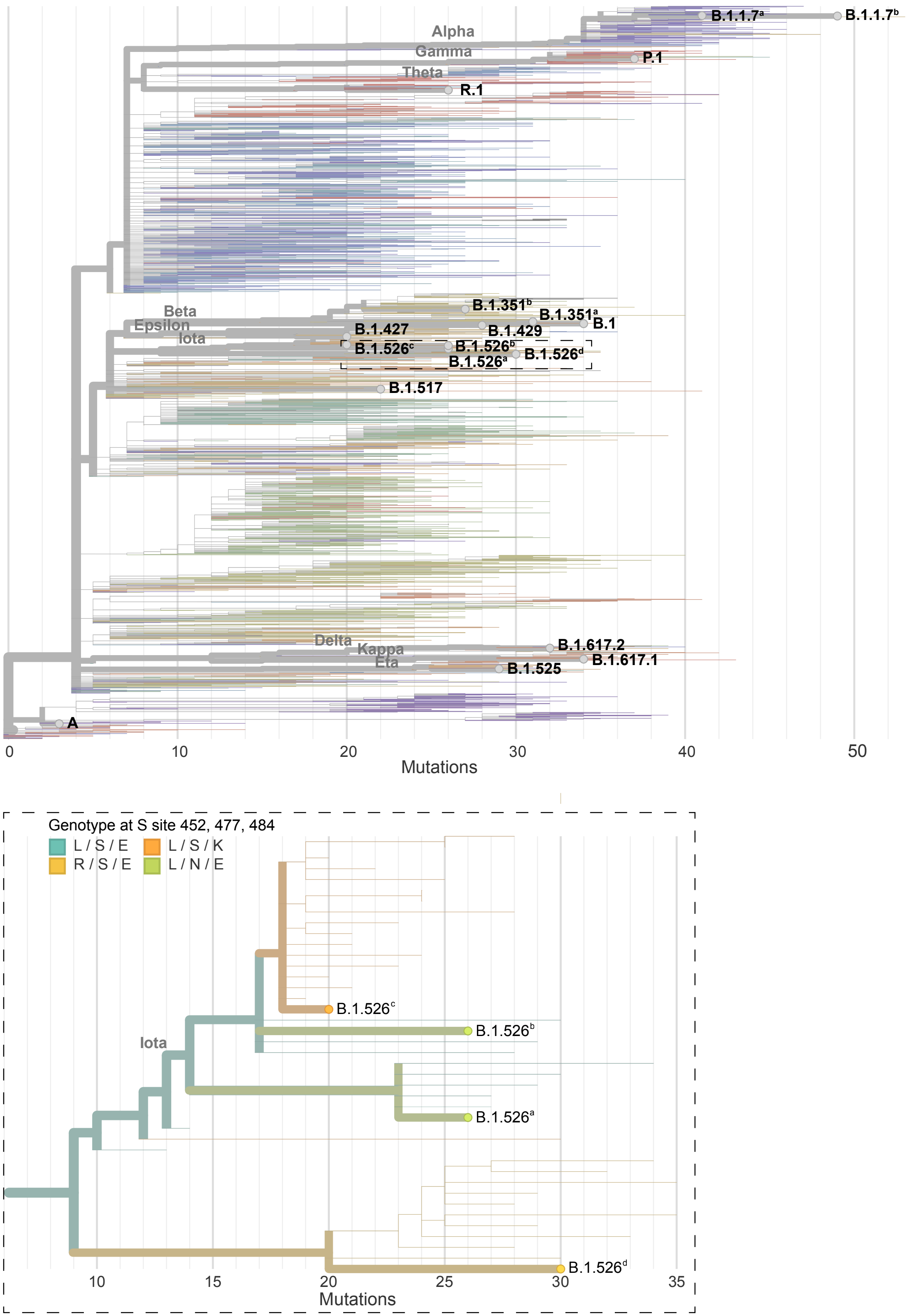

Alpha
Gamma
Theta
B.1.1.7[a]
B.1.1.7[b]
P.1
R.1
Beta
Epsilon
Iota
B.1.351[b]
B.1.351[a]
B.1
B.1.427
B.1.429
B.1.526[c]
B.1.526[b]
B.1.526[d]
B.1.526[a]
B.1.517
Delta
Kappa
Eta
B.1.617.2
B.1.617.1
B.1.525
A
0
10
20
30
40
50
Mutations
Genotype at S site 452, 477, 484
L / S / E
L / S / K
R / S / E
L / N / E
Iota
B.1.526[c]
B.1.526[b]
B.1.526[a]
B.1.526[d]
10
15
20
25
30
35
Mutations

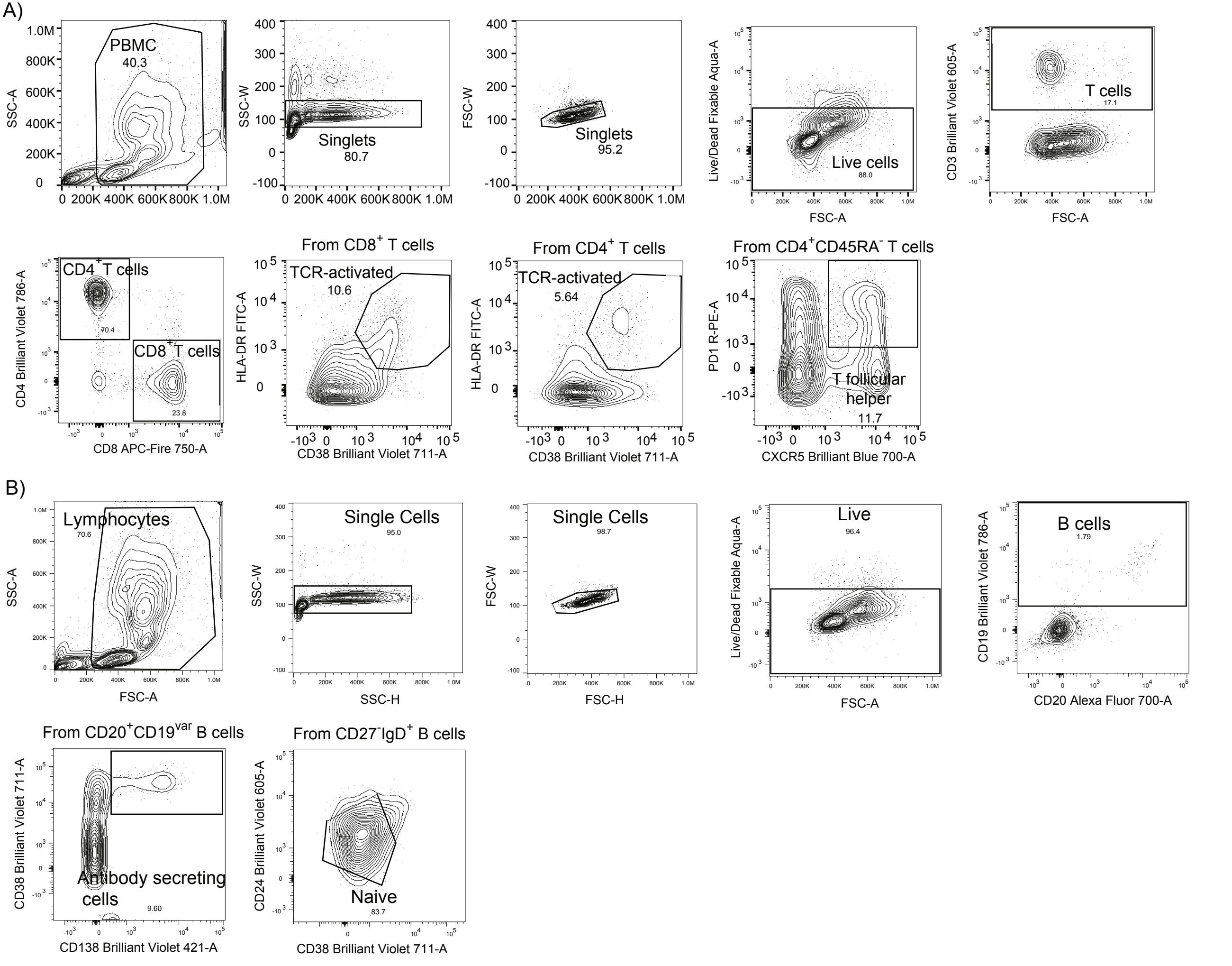
A)
PBMC
40.3
SSC-A
Singlets
80.7
SSC-W
Singlets
95.2
FSC-W
Live cells
88.0
Live/Dead Fixable Aqua-A
FSC-A
T cells
17.1
CD3 Brilliant Violet 605-A
FSC-A
CD4+ T cells
70.4
CD8+ T cells
23.8
CD4 Brilliant Violet 786-A
CD8 APC-Fire 750-A
From CD8+ T cells
TCR-activated
10.6
HLA-DR FITC-A
CD38 Brilliant Violet 711-A
From CD4+ T cells
TCR-activated
5.64
HLA-DR FITC-A
CD38 Brilliant Violet 711-A
From CD4+CD45RA- T cells
T follicular helper
11.7
PD1 R-PE-A
CXCR5 Brilliant Blue 700-A
B)
Lymphocytes
70.6
SSC-A
FSC-A
Single Cells
95.0
SSC-W
SSC-H
Single Cells
98.7
FSC-W
FSC-H
Live
96.4
Live/Dead Fixable Aqua-A
FSC-A
B cells
1.79
CD19 Brilliant Violet 786-A
CD20 Alexa Fluor 700-A
From CD20+CD19var B cells
Antibody secreting cells
9.60
CD38 Brilliant Violet 711-A
CD138 Brilliant Violet 421-A
From CD27-IgD+ B cells
Naive
83.7
CD24 Brilliant Violet 605-A
CD38 Brilliant Violet 711-A

| | | Vaccine | | Sex | | SeroStatus | |
|---|---|---|---|---|---|---|---|
| | Age (years) | Moderna | Pfizer | Female | Male | Negative | Positive |
| **Total Cohort %** | #DIV/0! | 80.56 | 19.44 | 80.56 | 19.44 | 50.0 | 50.0 |
| **Non- Previous exposed %** | 44.28 (1.833 ± 4.242) | 61.11 | 38.88 | 77.7 | 83.33 | | |
| **Previous exposed %** | 46.11 (1.833 ± 4.242) | 100.0 | 0.0 | 22.22 | 16.66 | | |

| Volunteers ID | Age range | Sex | SeroStatus | Vaccine |
|---|---|---|---|---|
| 1 | B | Female | Negative | Pfizer |
| 2 | B | Female | Negative | Pfizer |
| 3 | B | Female | Negative | Moderna |
| 4 | C | Female | Negative | Moderna |
| 5 | C | Female | Negative | Pfizer |
| 6 | B | Female | Negative | Pfizer |
| 7 | E | Female | Negative | Moderna |
| 8 | B | Female | Negative | Moderna |
| 9 | C | Male | Negative | Moderna |
| 10 | A | Female | Negative | Pfizer |
| 11 | B | Female | Negative | Moderna |
| 12 | B | Male | Negative | Moderna |
| 13 | D | Male | Negative | Moderna |
| 14 | A | Female | Negative | Moderna |
| 15 | D | Female | Negative | Pfizer |
| 16 | B | Female | Negative | Moderna |
| 17 | A | Female | Negative | Moderna |
| 18 | C | Male | Negative | Pfizer |
| 20 | D | Female | Positive | Moderna |
| 21 | A | Female | Positive | Moderna |
| 22 | D | Female | Positive | Moderna |
| 24 | E | Female | Positive | Moderna |
| 25 | C | Female | Positive | Moderna |
| 26 | B | Female | Positive | Moderna |
| 27 | C | Male | Positive | Moderna |
| 28 | D | Female | Positive | Moderna |
| 30 | A | Male | Positive | Moderna |
| 31 | A | Female | Positive | Moderna |
| 32 | C | Female | Positive | Moderna |
| 33 | A | Female | Positive | Moderna |
| 34 | A | Female | Positive | Moderna |
| 35 | C | Female | Positive | Moderna |
| 36 | D | Female | pOsitive | Moderna |
| 37 | D | Female | Positive | Moderna |
| 38 | D | Female | Positive | Moderna |
| 39 | A | Male | Positive | Moderna |

| Age Range | |
|---|---|
| A | 26-35 |
| B | 36-45 |
| C | 46-55 |
| D | 56-65 |
| E | 66-75 |

| **Lineage**<br>GenBank<br>Accession | **A**<br>MZ468053 | **B.1.526**[a]<br>MZ467323 | **B.1.526**[b]<br>MZ467333 | **B.1.1.7**[a]<br>MZ202178 | **B.1.517**<br>MZ468008 | **B.1.526**[c]<br>MZ201303 | **B.1.617.2**<br>MZ468047 | **R.1**<br>MZ467697 | **B.1.427**<br>MZ467318 | **B.1.526**[d]<br>MZ467322 | **B.1.429**<br>MZ467319 | **B.1.525**<br>MZ467313 | **B.1.617.1**<br>MZ468046 | **P.1**<br>MZ202306 | **B.1**<br>MZ467989 | **B.1.1.7**[b]<br>MZ467988 | **B.1.351**[a]<br>MZ468007 | **B.1.351**[b]<br>MZ202314 |
|---|---|---|---|---|---|---|---|---|---|---|---|---|---|---|---|---|---|---|
| E | | | | | | | V62F | | | | | L21F | | | | | P71L | P71L |
| M | | | | | | | I82T | F28L<br>T175M | | | | I82T | I82S | | | | | |
| N | | P13L<br>S202R<br>A414V | P199L<br>M234I | D3L<br>R203K<br>G204R<br>S235F | S206F | P199L<br>M234I | D63G<br>R203M<br>D377Y | S187L<br>R203K<br>G204R<br>Q418H | T205I<br>T362I | T205I<br>M234I | T205I<br>M234I | M1-<br>S2M<br>D3Y<br>A12G<br>T205I | R203M<br>D377Y | P80R<br>R203K<br>G204R | D3Y<br>T205I | D3L<br>R203K<br>G204R<br>S235F | T205I | T205I |
| ORF1a | | T265I<br>T1840I<br>G1946S<br>L3201P<br>S3675-<br>G3676-<br>F3677- | T265I<br>A1352V<br>L3201P<br>P3504S<br>S3675-<br>G3676-<br>F3677- | T1001I<br>P1213L<br>A1708D<br>I2230T<br>M2259I<br>S3675-<br>G3676-<br>F3677- | T265I<br>A541S<br>G989V<br>H1580Y | T265I<br>T2977I<br>L3201P<br>S3675-<br>G3676-<br>F3677- | P309L<br>A405V<br>P1640L<br>A3209V<br>V3718A | | T265I<br>S3158T | T265I<br>T2087I<br>K3162E<br>L3201P<br>A3209V<br>P3359S<br>S3675-<br>G3676-<br>F3677-<br>V3847I<br>L4126F | T265I<br>I4205V | T2007I<br>L2609I<br>S3675-<br>G3676-<br>F3677- | T1567I<br>T3646A | S1188L<br>K1795Q<br>G2941S<br>S3675-<br>G3676-<br>F3677- | G150S<br>T265I<br>V649F<br>T708I<br>A1049V<br>T1854I<br>K2497N<br>R3542C<br>M4375T | E913D<br>T1001I<br>A1306T<br>A1708D<br>P2046L<br>I2230T<br>M2259I<br>S3675-<br>G3676-<br>F3677-<br>L3736F<br>L3829F | T265I<br>T333M<br>Y1598C<br>K1655N<br>E1843D<br>T2174I<br>K3353R<br>S3675-<br>G3676-<br>F3677-<br>T4065I | T265I<br>K1655N<br>K3353R<br>S3675-<br>G3676-<br>F3677- |
| ORF1b | | P314L<br>Q1011H | P314L<br>Q1011H | P218L<br>P314L<br>A1432V | P314L<br>D1506N<br>P2633S | P314L<br>Q1011H<br>R1078C | P314L<br>G662S<br>P1000L<br>P1570L | P314L<br>G814C<br>G1362R<br>P1936H | P314L<br>P976L<br>D1183Y | P314L | P314L<br>D1183Y<br>G2436C | P314F | P314L<br>G1129C<br>A1291S<br>M1352I<br>K2310R<br>S2312A | P314L<br>A1219S<br>E1264D | T132I<br>P314L | P218L<br>P314L<br>T1511I | P314L<br>T1050I<br>Y2608H | P314L |
| ORF3a | | P42L<br>Q57H | P42L<br>Q57H | | Q57H<br>D210E | P42L<br>Q57H | S26L | | Q57H | P42L<br>Q57H | Q57H | S92L | S26L | Q57H<br>S253P | Q57H<br>P104S<br>S171L | | Q57H<br>S171L | Q57H<br>W131L<br>S171L |
| ORF6 | | | | | | | | | | | | M1-<br>F2M | | | | | | |
| ORF7a | | L116F | | | | | V82A<br>L116F<br>T120I | | | P34S | | A105S | N43Y<br>V82A | | | | | V93F |
| ORF8 | L84S | T11I | T11I | Q27*<br>R52I<br>K68*<br>Y73C | E59* | T11I | D119-<br>F120- | | | T11I<br>P36S<br>A51S | V100L | | | E92K | E106D | Q27*<br>R52I<br>K68*<br>Y73C | I121L | R115L |
| ORF9b | | P10S | T83I | | | | T60A | | | | | H9D | | Q77E | | | | |
| S | | L5F<br>T95I<br>D253G<br>S477N<br>D614G<br>Q957R | L5F<br>T95I<br>D253G<br>S477N<br>D614G<br>A701V | H69-<br>V70-<br>Y144-<br>N501Y<br>A570D<br>D614G<br>P681H<br>T716I<br>S982A<br>D1118H | N501T<br>Q613H<br>D614G<br>G639V | L5F<br>T95I<br>D253G<br>E484K<br>D614G<br>A701V | T19R<br>E156-<br>F157-<br>R158G<br>L452R<br>T478K<br>D614G<br>P681R<br>D950N<br>L1141W | W152L<br>E484K<br>K558N<br>D614G<br>G769V | S13I<br>W152C<br>L452R<br>D614G | D80G<br>Y144-<br>F157S<br>L452R<br>D614G<br>T791I<br>T859N<br>D950H | S13I<br>W152C<br>L452R<br>D614G | H69-<br>V70-<br>Y144-<br>Q52R<br>A67V<br>E484K<br>D614G<br>Q677H<br>F888L | T95I<br>G142D<br>E154K<br>L452R<br>E484Q<br>D614G<br>P681R<br>Q1071H | L18F<br>T20N<br>P26S<br>D138Y<br>R190S<br>K417T<br>E484K<br>N501Y<br>D614G<br>H655Y<br>T1027I<br>V1176F | E484K<br>N501T<br>D614G | H69-<br>V70-<br>Y144-<br>E484K<br>N501Y<br>A570D<br>D614G<br>P681H<br>T716I<br>S982A<br>D1118H | D80A<br>D215G<br>L241-<br>L242-<br>A243-<br>K417N<br>E484K<br>N501Y<br>D614G<br>A701V | L18F<br>D80A<br>D215G<br>L241-<br>L242-<br>A243-<br>K417N<br>E484K<br>N501Y<br>D614G<br>Q677H<br>R682W<br>A701V |

Abbreviations: E = envelope, M = membrane, N = nucleocapsid, ORF = open reading frame, S = spike.
